# Non-ablative stereotactic radiosurgery for subgenual cingulate neuromodulation in treatment-resistant depression: a randomized dose-seeking pilot trial

**DOI:** 10.64898/2026.08.13.26360283

**Authors:** Yingying Zhao, Yang Bai, Aihong Yu, Xiao Jin, Zhenxiang Zang, Fangcheng Zou, Qingyang Ma, Bin Wang, Xuequan Zhu, Zhi Yang, Hailun Hang, Yun Wang, Jinyuan Wang, Chengcheng Wang, Xiaoliang Liu, Yuxuan Xu, Qin Qin, Guangqiang Sun, Yuting Wang, Baolin Qu, Jianning Zhang, Ling Zhang, Hemmings Wu, John R. Adler, Longsheng Pan, Gang Wang

## Abstract

The subgenual anterior cingulate cortex (sgACC) is a key node in treatment-resistant depression (TRD), but precise non-invasive neuromodulation of this target is challenging. Preclinical studies of non-ablative stereotactic radiosurgery (SRS) have shown neuromodulatory (“radiomodulation”) effects. In this single-center, double-masked, randomized, dose-seeking pilot trial, nine adults with TRD were randomly assigned to bilateral sgACC radiomodulation at a dose of either 15, 20, or 25 Gy per hemispheric target. Primary endpoints were safety and feasibility; the efficacy endpoint was week-4 change in the Montgomery–Åsberg Depression Rating Scale (MADRS). Both primary endpoints were met: the only treatment-related adverse event was transient grade 1 dizziness, with no structural MRI abnormality through week 12. Mean MADRS fell from 33.0 to 17.0 (48.5% reduction); 67% responded and 44% remitted, with benefit sustained to week 12. Resting-state fMRI revealed regional connectivity changes correlating with clinical improvement, with tractography showing streamline counts differing by response status. These first-in-human findings support a larger randomized controlled trial of sgACC radiomodulation for TRD. ClinicalTrial.gov registration: <u>NCT07274917</u>.

## Main

Approximately 30% of adults with major depressive disorder (MDD) fail to remit after multiple antidepressant trials, a state termed treatment-resistant depression (TRD)^1,2^. Current interventions for TRD have substantial limitations: electroconvulsive therapy is associated with cognitive impairment; repetitive transcranial magnetic stimulation requires a prolonged treatment course; intranasal esketamine necessitates repeated dosing and is frequently accompanied by dissociative adverse effects; and circuit-based surgical therapies, such as vagus nerve stimulation and deep brain stimulation (DBS), entail perioperative risk and require hardware implantation^3–6^. The subgenual anterior cingulate cortex (sgACC) is a convergent depressogenic node whose dysregulated activity tracks symptom severity^7–9^. Prior lesioning approaches to this circuit, including both radiofrequency-based and high-dose stereotactic radiosurgery (SRS), the latter of which relies on focal single-fraction doses well above the radionecrotic threshold, permanently disconnect dysfunctional pathways^10,11^. With TRD, as with all circuit-based disorders of the brain, the search for better methods of neuromodulation is ongoing^12^.

At the highest level, the ideal neuromodulatory technique for TRD would need to be efficacious, durable, non-invasive and non-destructive, while enabling highly individualized and precise spatial targeting of the implicated brain region. Nearly a century of experience using ionizing radiation to treat conditions in and around the brain provides an enormous basis for establishing its relative safety. Past studies suggest that low doses of radiation delivered to small brain volumes via SRS are largely repairable and therefore without any clinically significant CNS effects^10,11,13^. However, numerous anecdotal observations in the field of SRS suggest that low-dose, non-ablative ionizing radiation can manifest meaningful, durable, and even circuit-wide neuromodulatory effects, via a mechanism termed radiomodulation^14–19^. While the underlying action mechanism remains a work in progress, we previously hypothesized that radiomodulation might provide clinically significant neuromodulation for selected intractable circuit-based mental illnesses^19^. With this objective in mind, and the above rationale, we set out to investigate the first-in-human potential of radiomodulation in a small cohort of patients with intractable functional brain disorders. While radiomodulation could theoretically benefit a range of disorders, we selected TRD because its underlying circuitry is comparatively well mapped^12,20^, converging on the sgACC (Brodmann area 25) as a focal node — a target whose modulation, notably via DBS, has already shown clinical benefit in TRD patients^21^.

In the present trial, the sgACC was irradiated with low-dose non-ablative SRS “radiomodulation”, the aim of which was to produce a graded, dose-dependent therapeutic effect. The radiation dose was deliberately set roughly an order of magnitude below that used to induce radionecrotic lesions (i.e., 140–180 Gy for capsulotomy)^14,16^. This first-in-human pilot study was designed to weigh the harms and benefits of sgACC radiomodulation in TRD. The primary objective of this first-in-human pilot study was to evaluate the safety and tolerability of bilateral radiomodulation targeted at the sgACC across escalating doses (15, 20, and 25 Gy per hemisphere), including dose-dependent adverse events as well as any radiological or neurocognitive evidence of injury. The secondary objective was to obtain preliminary estimates of antidepressant efficacy and to characterize the dose-response relationship, thereby both defining the risk–benefit profile and identifying a dose suitable for further investigation. Here, we report the first-in-human experience with bilateral sgACC radiomodulation in TRD patients, conducted as a single-center, double-masked, randomized, dose-seeking pilot trial (ClinicalTrials.gov ID: NCT07274917, https://clinicaltrials.gov/study/NCT07274917).

## Results

### Study participants: baseline characteristics

Between January and March 2026, nine TRD patients, from a total of eleven evaluated, were enrolled in the present investigation from the depression treatment center at Beijing Anding Hospital (Fig. 1); patient R003 was excluded due to an abnormal MRI screening, and patient R006 withdrew consent shortly after enrollment. The last enrolled patient completed the final follow-up visit on June 30, 2026. The treated cohort consisted of seven men and two women having a mean (± s.d.) age of 31.4 ± 8.7 years (range, 19–44). Patient age at first depressive episode averaged 23.6 ± 11.6 years with a current illness duration averaging 8.2 ± 7.7 years; three patients were in their third depressive episode, six patients in their second, and the mean number of episodes was 2.3 ± 0.5. At baseline, the cohort’s mean Montgomery-Åsberg Depression Rating Scale (MADRS) score was 33.0, the Hamilton Anxiety Rating Scale (HAM-A) score was 24.8, and the self-reported Inventory of Depressive Symptomatology (IDS-SR) score was 41.2. Baseline cognitive performance was within the expected range for this population (mean Digit Span Test 14.8; mean Digit Symbol Substitution Test 57.0), with an average self-reported Perceived Deficits Questionnaire–Depression (PDQ-D) of 40.3, indicating clinically meaningful perceived cognitive impairment. All patients continued their pre-existing concomitant medication regimens throughout the trial. Upon enrollment, each patient was randomly assigned to receive bilateral sgACC stereotactic radiomodulation using a dose of either 15 Gy (*n* = 3; R002, R004, R010), 20 Gy (*n* = 3; R007, R008, R009) or 25 Gy (*n* = 3; R001, R005, R011) per hemispheric target (Extended Data Figs. 1 and 2). Individual baseline characteristics, dose assignment and baseline severity are summarized in Table 1.

**Fig. 1.**
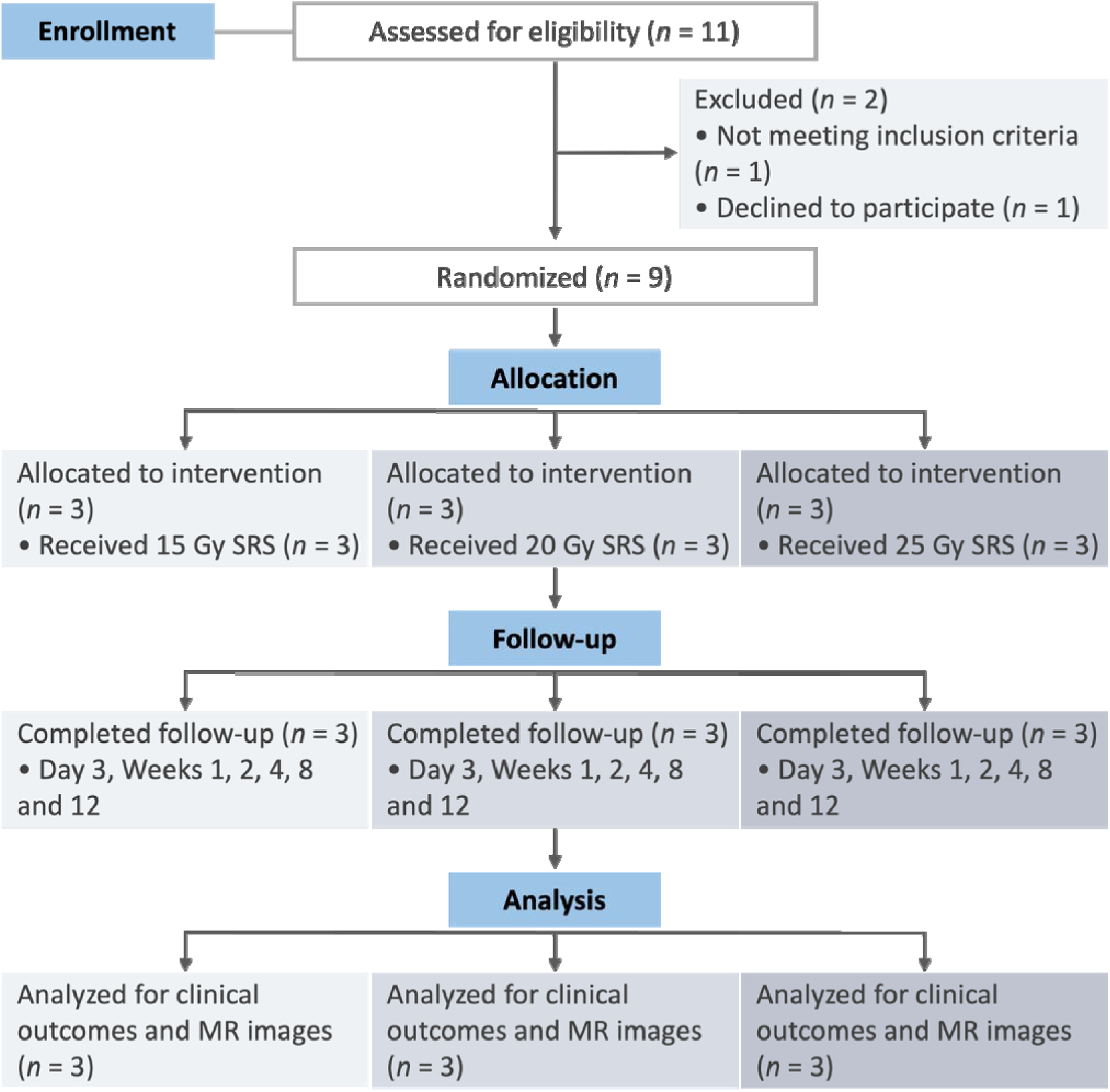
Participant flow through screening, enrollment, randomization, treatment and follow-up. Eleven participants were assessed for eligibility; two were excluded (one ineligible, one declining to participate). Nine participants completed baseline assessment (clinical, MRI and magnetoencephalography (MEG)), after which each was randomly assigned 1:1:1 to 15 Gy (*n* = 3), 20 Gy (*n* = 3) or 25 Gy (*n* = 3) per hemispheric target. All nine patients underwent sgACC radiomodulation delivered bilaterally over two consecutive days and were then followed through six post-SRS visits (day 3 and weeks 1, 2, 4, 8 and 12), each of which entailed rating scale, MRI and MEG.

**Table 1.** Baseline characteristics, dose assignment, and change of MADRS total score of the nine enrolled participants after treatment with SRS.

| ID | Age | Sex | Onset<br>(yr) | Duration<br>(yr) | Episodes | Dose<br>(Gy) | MADRS<br>(baseline / week 4<br>(post-treatment) / week 8<br>(post-treatment) / week 12<br>(post-treatment)) |
| --- | --- | --- | --- | --- | --- | --- | --- |
| R001 | 31-35 | F | 11-15 | 20 | 3 | 25 | 44 / 33 / 32 / 31 |
| R002 | 31-35 | M | 16-20 | 18 | 3 | 15 | 34 / 25 / 24 / 28 |
| R004 | 21-25 | M | 16-20 | 3 | 3 | 15 | 25 / 10 / 9 / 10 |
| R005 | 31-35 | M | 16-20 | 16 | 2 | 25 | 36 / 31 / 26 / 26 |
| R007 | 36-40 | M | 36-40 | 2 | 2 | 20 | 28 / 5 / 3 / 2 |
| R008 | 16-20 | M | 11-15 | 4 | 2 | 20 | 35 / 9 / 11 / 13 |
| R009 | 41-45 | M | 41-45 | 2 | 2 | 20 | 29 / 13 / 16 / 18 |
| R010 | 36-40 | M | 36-40 | 1 | 2 | 15 | 32 / 10 / 5 / 4 |
| R011 | 21-25 | F | 11-15 | 8 | 2 | 25 | 34 / 17 / 9 / 20 |

F, female; M, male; MADRS, Montgomery–Åsberg Depression Rating Scale.

### Safety and tolerability

All nine patients completed both treatment sessions and the prespecified 12–week post–treatment assessments, with no protocol–defined treatment discontinuations. The treatment was well tolerated across all three dose arms. The only treatment-related adverse event was transient dizziness reported by two patients immediately after their radiomodulation session; symptoms in both cases were self-limited, resolving without intervention over approximately fifteen minutes and were graded a Common Terminology Criteria for Adverse Events (CTCAE) grade 1 (mild) event. No other adverse incident was experienced. Through week 12 post-treatment, no abnormalities were observed on follow-up MRI in any patient (Fig. 2).

**Fig. 2.**
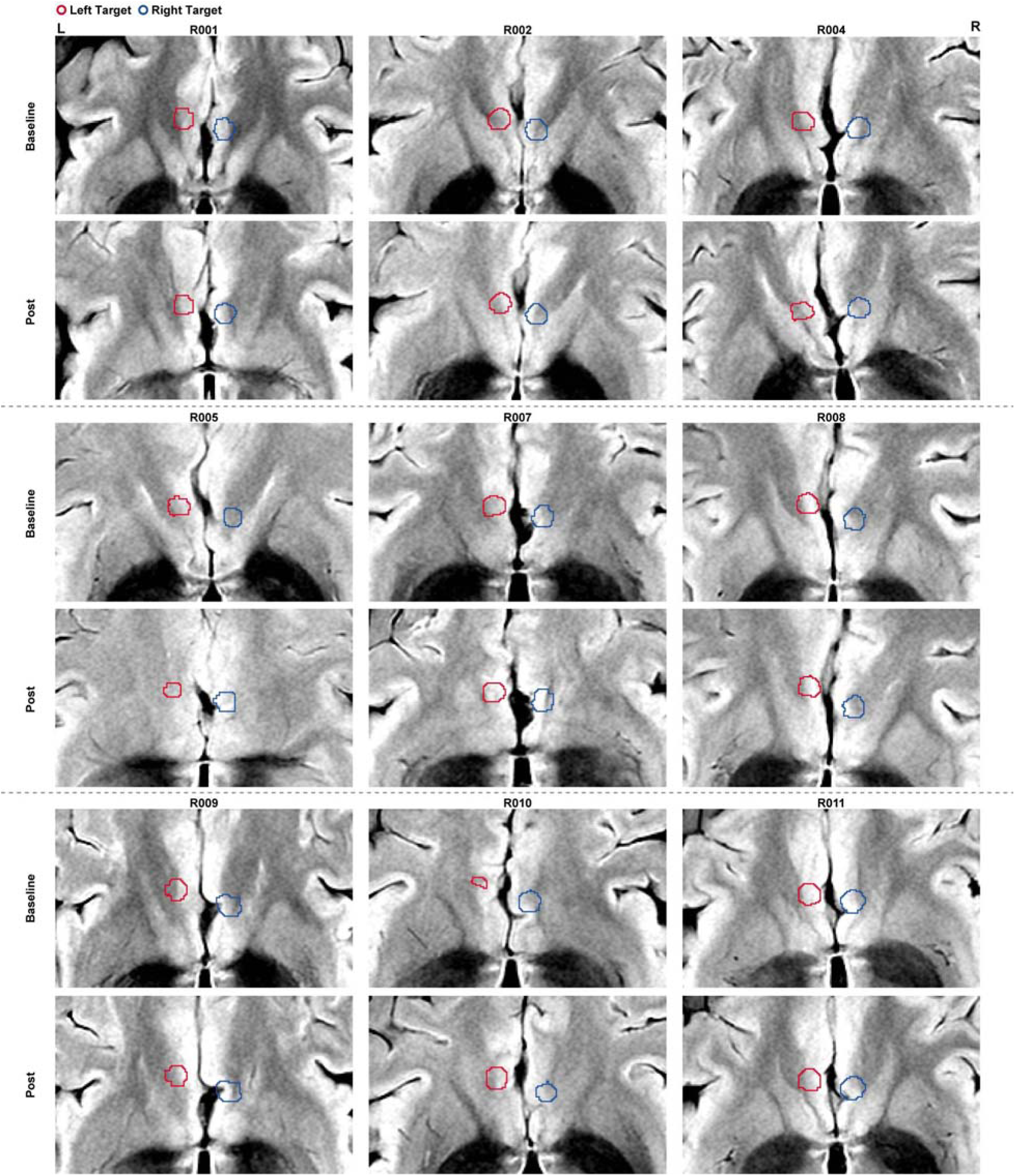
Serial FLAIR imaging of the bilateral sgACC targets before and after radiomodulation. Fluid-attenuated inversion recovery (FLAIR) images of the sgACC target for the nine patients. Red and blue contours indicate the approximate anatomical locations of the left and right sgACC treatment targets, respectively. Target labels were transformed into FLAIR space using Advanced Normalization Tools-derived registration and resampled separately onto a FLAIR-aligned intermediate grid with 1-mm through-plane spacing. No abnormalities were observed on MRI in any patient.

### Primary outcome: change in MADRS at week 4, with week 12 durability

The prespecified primary outcome for efficacy, summarized in Table 1, was the change in MADRS total score from baseline to week 4 after the second treatment session. Across the full cohort, the mean MADRS total score decreased from 33.0 ± 1.8 (mean ± s.e.m.) at baseline to 17.0 ± 3.4 at week 4; the group-mean percent change from baseline to week 4 was −48.5% (paired *t*(8) = −6.95, *P* < 0.001, FDR *q* < 0.001). Notably, a reduction was observed as early as the first post-treatment assessment (day 3), with mean MADRS scores (25.2) already lower than baseline and continuing to decrease through week 4 (Fig. 3a). At the prespecified week 12 durability time point, the cohort mean MADRS was 16.9, indicating durability of the antidepressant response. There was no evidence of relapse at the cohort level (Fig. 3a). Individual trajectories were heterogeneous: several patients showed large early reductions in MADRS that were sustained through week 12 (R004, R007, R008), whereas one patient (R001) showed only minimal, clinically insignificant improvement.

**Fig. 3.**
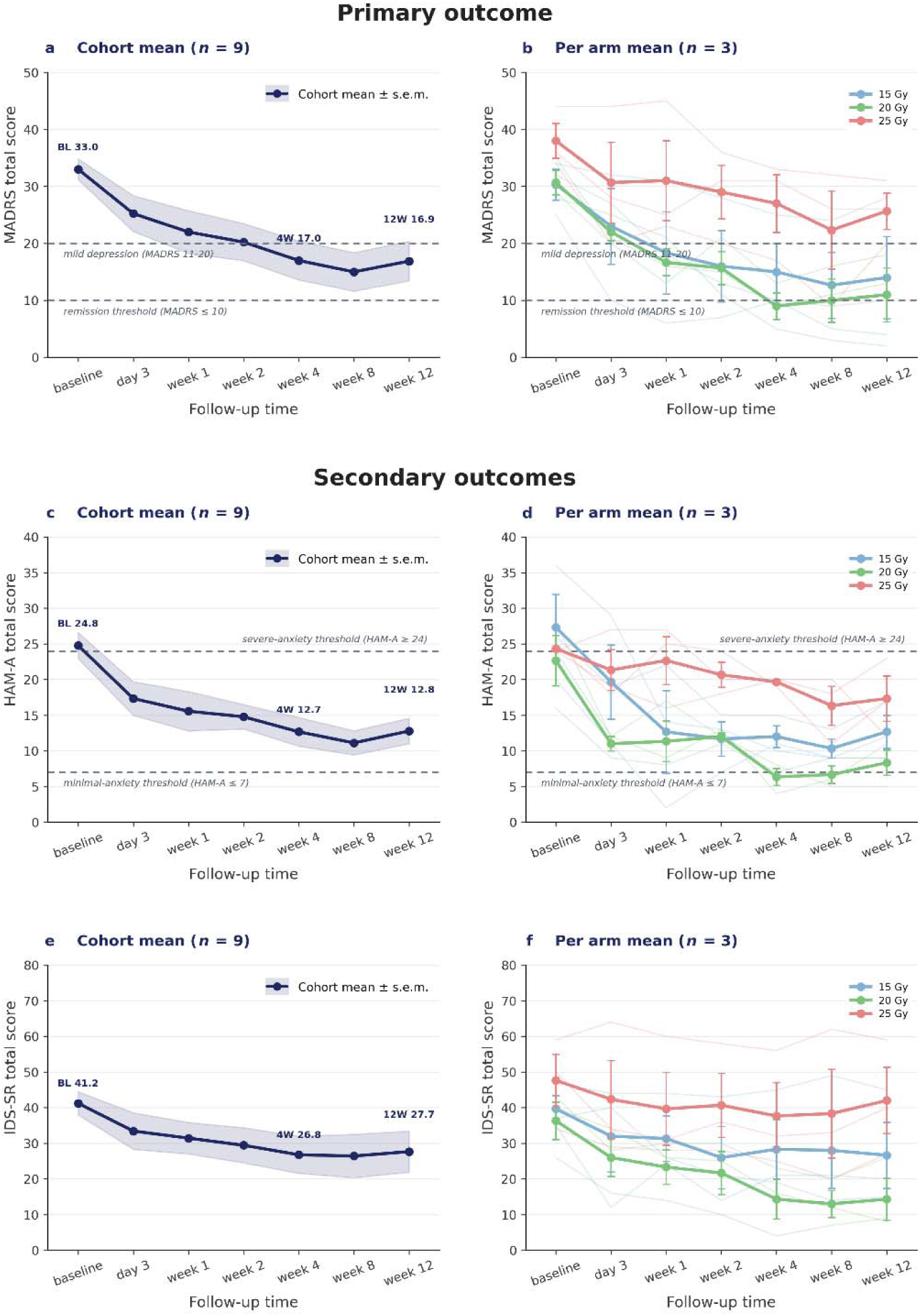
Primary and secondary efficacy outcomes (MADRS, HAM-A and IDS-SR) over the course of follow-up. **a**,**c**,**e**, Cohort mean ± s.e.m. (*n* = 9) at baseline, day 3, weeks 1, 2, 4, 8 and 12. For MADRS, the cohort mean percent change at week 4 was −48.5%; the cohort mean at week 12 was 16.9 (**a**). **b**,**d**,**f**, Per-arm means ± s.e.m. (*n* = 3 each) at 15 Gy (blue), 20 Gy (green) and 25 Gy (red) per hemispheric target, with faint lines showing individual subject trajectories. Dashed reference lines indicate the prespecified clinical thresholds: for MADRS, the remission threshold (≤ 10) and the upper bound of mild depression (11–20); for HAM-A, the minimal-anxiety threshold (≤ 7) and the severe-anxiety threshold (≥ 24).

Although the sample size (*n* = 3 per arm) precludes formal between-arm inference, the dose-stratified mean response showed the largest reduction in the 20 Gy arm, with a MADRS score of 9.0 at week 4 and 11.0 at week 12; a smaller mean reduction in the 15 Gy arm (15.0 at week 4, 14.0 at week 12); and the smallest reduction in the 25 Gy arm (27.0 at week 4, 25.7 at week 12; Fig. 3b). The week-4 to week-12 durability within every arm provides preliminary evidence that the early response to bilateral sgACC radiomodulation is seemingly sustained through at least three months post-treatment. At the prespecified week 4 primary endpoint, six of nine patients (66.7%; R004, R007, R008, R009, R010, R011) met the response criterion of a ≥ 50% reduction in the MADRS total score from baseline. Four of nine patients (44.4%; R004, R007, R008, R010) met the remission criterion of the MADRS score ≤ 10. Response and remission distributions across dose arms were uneven within the limits of *n* = 3 per arm: 2 of 3 responders and 2 of 3 remitters in the 15 Gy arm (R004, R010), 3 of 3 responders and 2 of 3 remitters in the 20 Gy arm (R007, R008), and 1 of 3 responders (R011) and 0 of 3 remitters in the 25 Gy arm. Between-arm effect sizes for the change in the MADRS score from baseline to week 4, along with MADRS response and remission rates at week 4, are provided in Extended Data Table 1.

### Secondary clinical outcomes

Clinician-rated anxiety symptoms, measured via the HAM-A mean total score, decreased in parallel with MADRS, from 24.8 at baseline to 12.7 at week 4, yielding a total reduction of 48.9% (paired *t*(8) = −4.56, *P* = 0.002, FDR *q* = 0.003; Fig. 3c). The dose-stratified pattern mirrored that on the MADRS: the largest mean reduction at week 4 occurred in the 20 Gy arm (6.3), with the arm mean below the minimal-anxiety threshold (HAM-A ≤ 7); the 15 Gy arm showed an intermediate reduction (12.0) and the 25 Gy arm the smallest (19.7; Fig. 3d). At the week 12 durability assessment, the cohort mean HAM-A was 12.8, essentially unchanged from week 4; within-arm values were stable from week 4 to week 12, indicating durability of the anxiolytic effect at the cohort level and within each dose arm. Self-rated depression on the Inventory of Depressive Symptomatology–Self Report (IDS-SR) decreased from a baseline mean of 41.2 to 26.8 at week 4 (group-mean reduction 35.03%; paired *t*(8) = −5.37, *P* < 0.001, FDR *q* = 0.001; Fig. 3e). The dose-stratified pattern also paralleled that observed on the MADRS: the largest mean reduction at week 4 occurred in the 20 Gy arm (22.0), while the 15 Gy arm showed an intermediate reduction (11.3) and the 25 Gy arm the smallest (10.0; Fig. 3f).

Cognitive assessment (DST, DSST and PDQ-D) results are provided in Extended Data Fig. 3. Clinician-administered cognitive performance improved on all measures: mean Digit Span Test (DST) scores improved from 14.8 to 16.3 (group-mean improvement 10.5%; paired *t*(8) = 3.09, *P* = 0.015, FDR *q* = 0.018; Extended Data Figs. 3a and 3b), and mean Digit Symbol Substitution Test (DSST) scores improved from 57.0 to 72.1 (paired *t*(8) = 5.31, *P* < 0.001, FDR *q* = 0.001; group-mean improvement 26.5%; Extended Data Figs. 3c and 3d). Self-rated perceived cognitive deficits on the Perceived Deficits Questionnaire–Depression (PDQ-D) decreased from a baseline mean of 40.3 to 28.7 at week 4 (group-mean reduction 28.9%; paired *t*(8) = −2.69, *P* = 0.027, FDR *q* = 0.027; Extended Data Fig. 3e); the dose-stratified means showed the largest reduction in the 15 Gy arm (Extended Data Fig. 3f). Between-arm effect sizes for all clinical outcomes from baseline to week 4 are reported in Extended Data Table 1a.

### Effects of time and illness duration on outcome scores over 12 weeks

Linear mixed-effects models revealed that time significantly predicted scores on all six measures after FDR correction: MADRS (*F*(6) = 62.43, FDR *q* < 0.001), HAM-A (*F*(6) = 41.75, FDR *q* < 0.001), IDS-SR (*F*(6) = 38.46, FDR *q* < 0.001), DST (*F*(6) = 12.69, FDR *q* = 0.048), DSST (*F*(6) = 52.85, FDR *q* < 0.001), and PDQ-D (*F*(6) = 20.02, FDR *q* = 0.003; detailed per-scale statistics in Extended Data Table 2). Longer illness duration was associated with persistently higher (worse) scores on the MADRS (β = 1.08, FDR *q* < 0.001), HAM-A (β = 0.46, FDR *q* < 0.001), IDS-SR (β = 1.70, FDR *q* < 0.001), and PDQ-D (β = 2.01, FDR *q* = 0.005), but was not associated with changes in objective cognitive performance on the DST (β = 0.03, FDR *q* = 0.892) or DSST (β = −0.09, FDR *q* = 0.892; Extended Data Fig. 4).

### Functional connectivity changes after sgACC radiomodulation

Because radiomodulation is hypothesized to act by modulating circuit function rather than by ablating tissue, we asked whether the observed clinical improvement was accompanied by measurable changes in sgACC connectivity. Using seed-based resting-state functional connectivity (FC) with the left and right sgACC as seeds, we examined treatment-related changes in connectivity and their relationship to symptom improvement. For the right sgACC (R.sgACC) seed, connectivity increased significantly with the medial prefrontal cortex (mPFC; *P*_FWE_ < 0.05, cluster size = 50 voxels; Figs. 4a and 4b). The change in R.sgACC–mPFC connectivity was negatively correlated with the MADRS reduction (*r* = −0.611, *P* = 0.081; Fig. 4c) and HAM-A reduction (*r* = −0.823, *P* = 0.006; Fig. 4d). For the left sgACC (L.sgACC) seed, connectivity with the left precentral gyrus (L.PreCG) was significantly reduced following radiomodulation (*P*_FWE_ < 0.05, cluster size = 27 voxels; Figs. 4a and 4e). The change in L.sgACC–L.PreCG connectivity was positively correlated with the MADRS reduction (*r* = 0.580, *P* = 0.102; Fig. 4f) and HAM-A reduction (*r* = 0.654, *P* = 0.056; Fig. 4g). Mirroring the significant findings, the R.sgACC showed a non-significant decrease in FC with the PreCG (Extended Data Fig. 5a) and the L.sgACC a non-significant increase in FC with the mPFC (Extended Data Fig. 5b), suggesting a broadly symmetric bilateral effect.

**Fig. 4.**
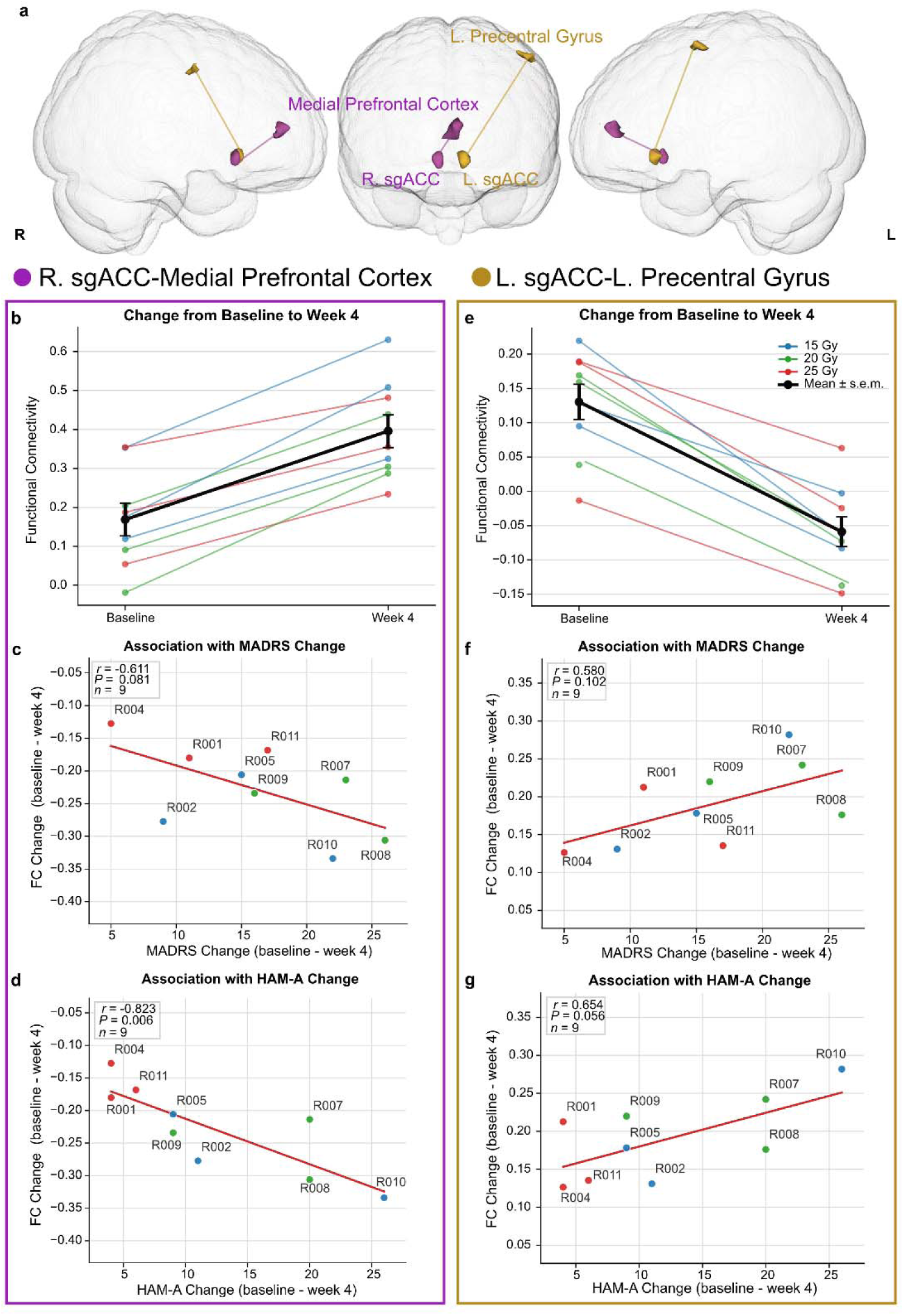
sgACC target-based functional connectivity (FC) changes following radiomodulation in TRD. Using the group-level union of individual sgACC stimulation targets as a seed, voxel-wise target-based FC was compared before and after radiomodulation (*n* = 9; cluster-level *P*_FWE_ < 0.05). **a**, Following bilateral sgACC radiomodulation, functional connectivity changed significantly between the right sgACC and the medial prefrontal cortex (mPFC), and between the left sgACC and the left precentral gyrus. **b**, Right sgACC (R.sgACC) FC was significantly *increased* with the mPFC (cluster size = 50 voxels); individual lines are colored by radiation dose (15 Gy, blue; 20 Gy, green; 25 Gy, red), and the bold black line denotes the group mean ± s.e.m. **c**,**d**, Association between the R.sgACC-mPFC FC change and MADRS/HAM-A reduction from baseline to week 4 (*n* = 9); each point represents one patient, colored by radiation dose (15 Gy, blue; 20 Gy, green; 25 Gy, red), with the fitted regression line shown. **e**, Left sgACC (L.sgACC) FC was significantly *decreased* with the left precentral gyrus (L.PreCG; cluster size = 27 voxels); individual lines are colored by radiation dose (15 Gy, blue; 20 Gy, green; 25 Gy, red), and the bold black line denotes the group mean ± s.e.m. **f**,**g**, Association between the L.sgACC-L.PreCG FC change and MADRS/HAM-A reduction from baseline to week 4 (*n* = 9); each point represents one patient, colored by radiation dose (15 Gy, blue; 20 Gy, green; 25 Gy, red), with the fitted regression line shown.

### sgACC target structural connectivity and week 4 clinical response

Having observed variability in clinical response, we next asked whether baseline anatomy could account for it. Because the therapeutic effect of sgACC neuromodulation is thought to depend on white-matter connections, we used diffusion tractography to test whether pre-treatment structural connectivity of the target area predicted clinical response at week 4. Patients were stratified by clinical response status at week 4, defined as responders (≥ 50% reduction in the MADRS total score from baseline) and non-responders (< 50% reduction). Compared with non-responders, responders showed significantly greater left sgACC-seeded streamline counts traversing the bilateral cingulum bundle (mean ± s.e.m.: responders, 2076.2 ± 367.5; non-responders, 909.7 ± 237.4; *t* = 2.67, *P* = 0.032, Hedges’ *g* = 1.31; Figs. 5a and 5b). Responders also showed higher counts of bilateral sgACC-seeded streamlines terminating in the mPFC than non-responders (mean ± s.e.m.: responders, 570.5 ± 180.6; non-responders, 209.7 ± 49.0), although this difference did not reach statistical significance (*t* = 1.93, *P* = 0.105, Hedges’ *g* = 0.85; Figs. 5c and 5d).

**Fig. 5.**
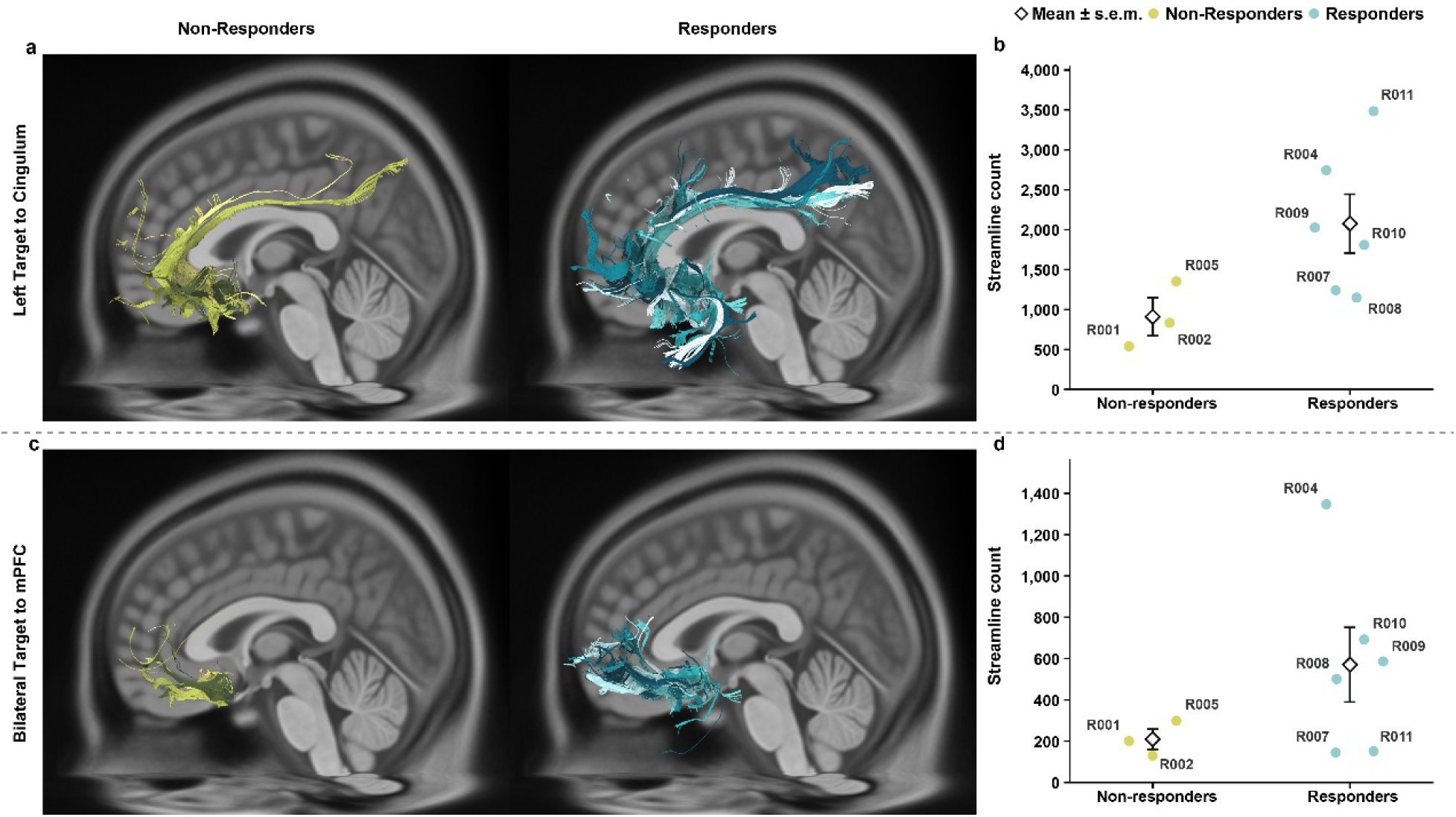
sgACC-seeded tractography by week 4 clinical response status. **a**, Standard-space overlays of individual tractography results traversing the bilateral cingulum bundle for non-responders and responders. **b**, Individual streamline counts and group mean ± s.e.m. for non-responders and responders, showing significantly greater streamline counts in responders (*P* = 0.032). **c**, Standard-space overlays of individual tractography results terminating in the medial prefrontal cortex (mPFC) for non-responders and responders. **d**, Individual streamline counts and group mean ± s.e.m. for non-responders and responders, showing higher counts in responders that did not reach significance (*P* = 0.105).

## Discussion

This dose-exploration study provides the first clinical evidence that bilateral low-dose stereotactic radiosurgery to the sgACC is feasible, well tolerated and associated with clinically meaningful antidepressant effects in patients with treatment-resistant depression. Using the ZAP-X gyroscopic platform at doses of 15–25 Gy, we observed a response in 6 of 9 patients (67%) and remission in 4 of 9 (44%) at the week-4 primary endpoint. Notably, no serious adverse events occurred in any of the nine patients: serial cognitive testing revealed no detectable deterioration, and no changes were observed on serial MR imaging—the latter contrasting with the longer-term MRI changes reported in the earlier 75 Gy series^10^ and consistent with the appreciably lower, sub-ablative doses used here. Clinical improvement was paralleled by neuroimaging changes consistent with modulation of sgACC-anchored circuitry, and the patients who responded most were those whose treatment volume encompassed more of the cingulum bundle projections. Although these findings are preliminary and warrant long–term surveillance, they provide initial support for the hypothesis that ionizing radiation at sub-ablative doses can exert sustained neuromodulatory effects in the human brain, and they provide a rationale for controlled investigation of radiosurgical neuromodulation as a noninvasive therapy for TRD and, potentially, for other circuit-based disorders.

The therapeutic armamentarium for TRD now includes a spectrum of neuromodulatory strategies that differ markedly in invasiveness, spatial precision, reversibility and evidentiary maturity; low-dose sgACC radiomodulation occupies a distinctive position among them. Repetitive transcranial magnetic stimulation (rTMS) is considered the least invasive option, and its accelerated, functional-connectivity-guided form has substantially shortened the treatment course and improved remission rates; however, rTMS delivers superficial cortical stimulation that reaches the sgACC only indirectly through cortico-cingulate connectivity, requires repeated sessions, and produces effects that typically wane without maintenance^22^. Vagus nerve stimulation (VNS) has shown durable benefit on response, functional and quality-of-life measures, but it entails surgical device implantation, indirect and diffuse afferent modulation, and a characteristically long latency to benefit that is measured in months^23^. Subcallosal cingulate deep brain stimulation (SCC-DBS) engages essentially the same circuit we targeted in this trial and, in long-term pooled cohorts, yields sustained response in many patients. However, SCC-DBS remains investigational, requires craniotomy, permanent hardware implantation and ongoing programming, and carries the attendant risks of infection, lead revision and device-related complications^24^. Magnetic resonance-guided focused ultrasound (MRgFUS) capsulotomy is incisionless like radiosurgery but is fundamentally ablative, creating a fixed thermal lesion of the anterior limb of the internal capsule, and its use in depression is confined to small early-phase series^25^. Against this backdrop, sub-ablative sgACC radiosurgery is uniquely noninvasive – requiring neither craniotomy, implanted hardware nor repeated procedural sessions – while delivering sub-millimetric, circuit-specific dosing directly to a validated depression node. Radiomodulation thus offers a potentially favorable balance of anatomical precision and low procedural burden, positioning it as a more scalable treatment option, pending validation against these comparators in controlled trials.

The only prior clinical precedent for radiosurgical targeting of the sgACC is the three-patient series reported by Solvason et al., in which the bilateral sgACC was treated at a much higher dose of 75 Gy for treatment-resistant bipolar depression; two of the three patients achieved a greater than 50% reduction in symptoms at 12 months^10^. Our results extend this early observation to unipolar TRD and, critically, show that a three- to five-fold lower, explicitly non-ablative dose can produce clinically meaningful antidepressant effects after a comparatively short latency. This dissociation between dose and effect is informative. Given that efficacy was observed at radiation levels well below the ablative threshold, tissue destruction is unlikely to be the mechanism underlying the antidepressant response. That interpretation is reinforced by the convergent findings of the companion study by Fan et al., which, together with our data, support a model in which the therapeutic action of radiomodulation arises from sub-ablative circuit modulation rather than from lesioning.

Although the precise pathways remain incompletely characterized, the connectivity changes we observed after sub-ablative radiomodulatory doses (15–25 Gy) map onto well-established models of sgACC circuit dysfunction in depression. We found that changes in L.sgACC–L.PreCG connectivity correlated positively with reductions in MADRS score, whereas changes in R.sgACC–mPFC connectivity correlated negatively. This pattern is consistent with the sgACC’s role as a pivotal node in mood regulation whose aberrant coupling to prefrontal and motor regions correlates with the pathophysiology of depression^26^. The positive L.sgACC–L.PreCG association plausibly reflects a restoration of emotional–motor integration: the precentral gyrus subserves motor planning and volitional action, and strengthening its coupling with the left sgACC may attenuate psychomotor retardation and facilitate goal-directed behavior, a core deficit in depression^27^. The negative R.sgACC–mPFC association, by contrast, fits the hyperconnectivity model of ruminative self-focus, in which overactive coupling between the sgACC and the self-referential mPFC sustains maladaptive rumination^28^; a treatment-related decrease in this connectivity would disengage a circuit that perpetuates negative affect, with larger decreases tracking greater symptom relief.

The integrity of sgACC–mPFC connectivity is central to the pathophysiology of depression, and its dysregulation is associated with impaired emotional regulation and behavioral control^29,30^.

In children with a history of preschool-onset MDD, for example, aberrant connectivity between the sgACC and the dorsomedial prefrontal cortex (dmPFC) correlates significantly with dysregulated emotional behavior^31^. Similarly, longitudinal studies in adolescents have demonstrated that changes in SGC-dmPFC connectivity are predictive of depressive symptom severity, highlighting the role of this circuit in the developmental emergence and persistence of the disorder^32^. The sgACC and mPFC are integral components of the default mode network (DMN), a system often characterized by hyperconnectivity in depressed populations^33^. This DMN-related circuitry is implicated in self-referential processing and the maintenance of depressive states^34^. Metabolic connectivity patterns underscore this, as the sgACC exhibits distinct lateralized connections with ventromedial prefrontal regions—areas associated with anhedonia and self-referential thought—in patients with refractory melancholic depression^34^. Because the sgACC is a central hub for these DMN-related processes, its interaction with the mPFC is considered a primary target for modulating the neural states that perpetuate depressive symptoms^33^. Functional connectivity between the sgACC and mPFC serves as a potential biomarker for predicting clinical response to various depression treatments^29,35^. In patients with TRD, baseline sgACC-dmPFC connectivity has been shown to correlate positively with clinical improvements following interventions^29^. Furthermore, neuromodulation therapies, particularly accelerated high-frequency repetitive transcranial magnetic stimulation (HF-rTMS), demonstrate efficacy by acutely adjusting these deregulated networks^35^. Successful rTMS treatment in TRD patients is often characterized by the strengthening of anti-correlations between the sgACC and the left superior medial prefrontal cortex, suggesting that restoring normative connectivity patterns is essential for clinical recovery^35^. That normalization of the same circuitry accompanies response across modalities lends external validity to the connectivity shifts we observed after radiomodulation. Together, these opposing correlations describe a double dissociation in which effective treatment simultaneously strengthens a left-lateralized circuit supporting motivated motor output and weakens a right-lateralized circuit underlying depressive rumination^29^.

Beyond these functional-connectivity findings, structural imaging pointed to the anatomical substrate through which radiomodulation may act. Our tractography analysis showed greater cingulum bundle streamline counts within the targeted sgACC in responders than in non-responders, suggesting that engagement of this tract may be associated with subsequent clinical improvement. Structural and functional disruptions in the cingulum bundle are linked to depression severity and treatment response. Changes in white matter integrity within this tract affect connectivity between limbic and cortical regions, influencing both symptom manifestation and therapeutic outcomes. Research utilizing diffusion tensor imaging (DTI) indicates that depression is characterized by reduced fractional anisotropy (FA) and axial diffusivity (AD), alongside increased radial diffusivity (RD) within both the dorsal and ventral segments of the cingulum bundle^36^. These white matter abnormalities are closely correlated with cumulative and current depression severity, suggesting that the integrity of this tract is a critical biomarker for the disorder. As a major white matter tract, the cingulum bundle facilitates essential connectivity between the anterior cingulate cortex (ACC), the hippocampus, and other limbic structures involved in affective regulation^37^. Functional damage to the cingulum bundle in the hippocampal region is associated with an increased predisposition to MDD and serves as a potential predictor of antidepressant efficacy. Furthermore, evidence suggests that the amount of cingulum bundle activation is a key factor in recovery for patients undergoing subcallosal cingulate deep brain stimulation (DBS)^38^. Because this bundle connects the subgenual cingulate cortex (sgACC) to wider networks, it is considered a primary target for DBS interventions in treatment-resistant depression^39^. The reliance on these pathways for interoceptive predictions indicates that disruptions in the cingulum bundle may undermine the brain’s ability to regulate affective states, thereby contributing to the clinical presentation of depression^40^.

There are several limitations to this study. First is the small pilot sample; with nine patients treated across three dose levels, it remains unknown whether these results will generalize. The absence of a sham comparator is a further limitation, as spontaneous fluctuation, regression to the mean and expectancy cannot be excluded as contributors to the observed response. It is also possible that the favorable safety profile reflects the limited follow-up available to date, since delayed radiation effects may emerge over a period of months to years and will require continued surveillance. Finally, targeting was based on anatomical landmarks, which may not capture individual functional-circuit variability; individualized tractography- or connectivity-guided targeting may refine both precision and efficacy in future work. The intent of this study was not to establish the efficacy of radiosurgical neuromodulation for treatment-resistant depression, the evidence for which will require a double-blind, randomized, sham-controlled and adequately powered trial. Rather, in this study we established proof-of-concept that ionizing radiation delivered at sub-ablative doses can produce sustained, circuit-level neuromodulatory effects in the human brain. This framework reframes stereotactic radiosurgery as a noninvasive tool for targeted circuit modulation and could advance the mechanistic understanding and treatment of a broad range of neuropsychiatric and other circuit-based disorders.

## Methods

### Study design and oversight

This prospective, randomized, parallel-group, open-label, double-masked exploratory trial – with patients and outcomes assessors masked to treatment assignment and the investigator unmasked – investigated stereotactic irradiation of the bilateral sgACC in adults with TRD. Conducted between January and June 2026, the study was a collaboration between Beijing Anding Hospital and Chinese PLA General Hospital. Before any patient recruitment, the trial protocol was approved by the institutional review boards and the independent ethics committees of both participating sites, subsequent to which each patient provided written informed consent.

### Participants

Eligible patients were outpatients or inpatients in the depression treatment center of Beijing Anding Hospital and aged 18 to 50 years (inclusive), of any sex, meeting DSM-5 diagnostic criteria for recurrent MDD without psychotic features. TRD was defined as a lack of clinically meaningful response (less than 50% improvement in depressive symptoms) to at least two adequate antidepressant trials during the current episode, documented using the Massachusetts General Hospital Antidepressant Treatment Response Questionnaire (MGH-ATRQ); an adequate trial was defined as sufficient dosage within the recommended therapeutic range and a duration of at least six weeks; the current episode needed to include at least one treatment failure. The 17-item Hamilton Depression Rating Scale (HAMD-17) total score was required to be ≥ 20 at screening. All patients maintained their existing baseline antidepressant regimen throughout the study.

Key exclusion criteria were any other current or prior major psychiatric disorder diagnosed according to DSM-5, such as schizophrenia, bipolar disorder, neurodevelopmental disorders, substance use disorder; clinically significant somatic, oncologic or severe neurological disease (including stroke, intracranial haemorrhage, raised intracranial pressure, space-occupying lesion, seizure history, cerebral aneurysm, Parkinson’s, Huntington’s, multiple sclerosis or severe head trauma with loss of consciousness); significant suicide risk (C-SSRS Suicidal Ideation Item 4 or 5 within six months, any Suicidal Behavior item within six months, or MADRS Item 10 ≥ 5); other neuromodulation (MECT, rTMS, tDCS, VNS or DBS) within three months, or any interventional trial within one month; non-response to MECT; prior psychosurgery; any prior radiotherapy, chemotherapy or immunotherapy or radiation/toxic occupational exposure; MRI contraindications; pregnancy, lactation or planned pregnancy within six months without effective contraception.

### Randomization and masking

Eligible participants were randomized in a 1:1:1 ratio to the 15 Gy, 20 Gy or 25 Gy dose group using a block randomization scheme (block size 3); the randomization sequence was generated using SAS software by an independent statistician not otherwise involved in trial conduct, and allocation was managed centrally. Once a participant was enrolled, the randomization administrator informed the treating investigator of the assigned dose group; the investigator was therefore not masked to dose allocation. As treatment planning and delivery required knowledge of the assigned dose, the radiation treatment team was likewise not masked. Participants were aware that they were receiving active radiotherapy but were not informed of their assigned dose group, and outcomes assessors remained masked to dose allocation throughout the study.

### Target volume delineation workflow

During the baseline period, patients sequentially underwent multimodal imaging examinations, including CT, contrast-enhanced MRI, functional magnetic resonance imaging (fMRI) and DTI to acquire high-resolution anatomical and functional data. Following the established planning workflow, the anatomical images were imported into MIM (MIM® Software 6.9.5 Build JB06-04) for image fusion. Key anatomical landmarks and structures were repeatedly compared during the fusion process to ensure accurate alignment across imaging modalities. Guided by the functional–anatomical principles and with particular reference to functional imaging cues, target delineation was performed on the fused images. A near-spherical volume with a diameter of 5 mm was used as the basic delineation unit. Bilateral sgACC targets were defined manually using the following anatomical workflow: on axial enhanced 3D T1-weighted MRI, the anterior commissure (AC) and posterior commissure (PC) were identified; on the sagittal T1-weighted MR image, the cingulate sulcus immediately below the genu of the corpus callosum was identified, a line was drawn from the genu of the corpus callosum to the AC, and the midpoint of this line was taken; on the coronal T1-weighted MR image, the coronal section corresponding to this midpoint was identified as the target point, which was then adjusted manually to avoid overlap with cerebral blood vessels. Simultaneously, protective contours were generated around adjacent critical structures and vessels traversing the target area to minimize irradiation of surrounding tissues. Once target delineation was complete, dose determination and treatment planning were performed, aiming to balance target coverage, dose conformity, and a steep dose gradient, thereby ensuring optimal target irradiation while maximizing patient safety.

### Intervention and quality assurance

Treatment was delivered using the ZAP-X gyroscopic radiosurgery robot (ZAP Surgical Systems, San Carlos, CA, USA) at the Department of Neurosurgery, the First Medical Center, Chinese PLA General Hospital. ZAP-X radiosurgery was delivered over two consecutive days; on each day, one hemispheric target received a single unilateral fraction of 15 Gy, 20 Gy or 25 Gy to the 80% isodose line according to the assigned arm, with the left hemisphere treated on day 1 and the right hemisphere on day 2, and each session lasting approximately 20 minutes. Real-time image guidance and intra-fractional motion monitoring were employed to maintain targeting accuracy of 0.5 mm, and the system automatically paused if the delivered dose deviated by more than 10% from the planned dose. Premedication with dexamethasone could be administered prophylactically at the treating physician’s discretion. All nine patients completed both treatment sessions per protocol.

### MRI data acquisition

MRI data were acquired at all scheduled time points for 9/9 patients; all datasets passed quality control (mean framewise displacement < 0.5 mm). Neuroimaging data were acquired at baseline and at 4 weeks and 12 weeks post-treatment at Beijing Anding Hospital using a 5.0 T MR scanner (uMR Jupiter, United Imaging Healthcare, Shanghai, China). The protocol comprised T1-weighted structural MRI, resting-state functional MRI and DTI. Acquisition parameters were as follows: (1) High-resolution T1-weighted imaging using a 3D gradient-echo sequence: TR = 9.2 ms, TE = 3.1 ms, FOV = 256 × 220 mm, 260 slices, slice thickness = 0.70 mm, FA = 9°. (2) Resting-state functional MRI: TR = 2000 ms, TE = 24 ms, FOV = 211 × 210 mm, acquisition matrix = 132 × 132, 90 slices, slice thickness = 1.6 mm, no slice gap, FA = 80°. (3) Arterial spin labelling: a 3D multi-post-labelling-delay pseudo-continuous ASL sequence, TR = 6000 ms, TE = 13.9 ms, acquisition matrix = 64 × 64, voxel size = 3.50 × 3.50 × 4.00 mm³, FOV = 224 × 224 mm, 32 slices, slice thickness = 4.00 mm, post-labelling delays of 500, 1000, 1500, 2000 and 2500 ms, with background suppression and a separate M0 calibration image. During scanning, patients were asked to keep their eyes closed, relax, not think intentionally, and not fall asleep. Routine clinical sequences and a contrast-enhanced MRI (baseline preparation period) were also acquired.

### Resting-state fMRI preprocessing and analysis

Resting-state fMRI data were preprocessed using SPM12 (version 7771; http://www.fil.ion.ucl.ac.uk/spm), MATLAB R2019a (MathWorks, Natick, MA, USA) and the Data Processing Assistant for Resting-State fMRI (DPARSFA 5.5; http://www.rfmri.org/DPARSF). The pipeline comprised removal of the first five volumes, slice-timing correction, head-motion correction, nuisance covariance regression, normalization of the functional images to the T1 image, resampling to 2 × 2 × 2 mm³, spatial smoothing with a 4 mm kernel, and band-pass filtering (0.01–0.1 Hz). Micro-head motion was quantified as the mean framewise displacement (FD); patients with a mean FD exceeding three interquartile ranges above the sample median, or with more than 2 mm maximum translation or 2° rotation about any axis in any volume, were to be excluded. On this basis, no patient was excluded from the final analysis.

SgACC target-based functional connectivity (FC) was computed to characterize brain function. Each patient’s individual stimulation target was delineated in native space and normalized to Montreal Neurological Institute (MNI) space; a group-level target region of interest (ROI) for each hemisphere was created as the union of all normalized individual targets, and this union ROI was used as a seed to compute the FC between the sgACC target and every other voxel across the whole brain.

To identify regions showing significant changes after stimulation, paired *t*-tests were used to compare each functional metric between the final baseline assessment and the 4-week follow-up. Whole-brain results were corrected for multiple comparisons using a voxel-wise threshold of *P* < 0.001 combined with a cluster-wise FWE threshold of *P*_FWE_ < 0.05. Finally, correlation analyses assessed the relationship between functional changes (baseline minus follow-up values) and clinical improvement at the 4-week follow-up, with significance set at *P* < 0.05.

### Outcomes

The primary efficacy outcome was the change in MADRS total score at four weeks after treatment, compared with baseline. Secondary efficacy and clinical outcomes, all assessed as a change from baseline at prespecified post-treatment time points, were the MADRS, the Hamilton Anxiety Rating Scale (HAM-A), the 30-item Inventory of Depressive Symptomatology–Self-Report (IDS-SR), the Digit Span Test (DST), the Digit Symbol Substitution Test (DSST) and the Perceived Deficits Questionnaire–Depression (PDQ-D). The Columbia–Suicide Severity Rating Scale (C-SSRS) was administered at eligibility screening and each follow-up visit as a categorical safety measure (see Participants) rather than as an analyzed efficacy outcome. The Clinical Global Impression–Severity (CGI-S) and Improvement (CGI-I) scales are single-item clinician-rated global measures collected at each visit for clinical characterization and were not treated as key outcomes for statistical analysis. The Immediate Mood Scaler-12 (IMS-12; including immediate intra-treatment assessments on days 1 and 2) is a momentary self-rated mood measure judged better suited to capturing the rapid-onset effects of interventions, such as ketamine, than the slower therapeutic trajectory expected of stereotactic radiomodulation, and its data were thus not analyzed in this study. Other prespecified outcomes were prespecified safety endpoints and an adverse event grading scheme at every follow-up visit.

### Sample size and statistical analysis

The estimated enrollment was nine patients (three per dose arm). Given the exploratory, dose-finding nature of the study, a formal sample-size calculation was not performed. The cohort size of three patients per dose arm is consistent with cohort sizes used in prior early-phase feasibility studies of stereotactic radiosurgery for psychiatric indications, including a study of bilateral subgenual cingulate (SGC) cortex radiosurgery for treatment-resistant bipolar depression (*n* = 3 total); that study, however, used a single fixed dose rather than a dose-escalation design^10^. The three-arm, three-patients-per-arm structure of the present study was selected as a pragmatic, feasibility-oriented approach to preliminary dose characterization across the 15–25 Gy range, rather than one derived from a formal statistical power or precision-based justification.

Clinical data were analyzed using IBM SPSS Statistics 26 and Python (version 3.10.12; pandas 2.3.3, statsmodels 0.14.6, matplotlib 3.10.9). Demographic characteristics and change scores from baseline are presented as means ± s.d. for continuous variables and frequencies (percentages) for categorical variables; group-level summaries of clinical and neuroimaging measures are presented as mean ± s.e.m. All nine patients received the full treatment protocol and completed all six follow-up assessments (day 3, weeks 1, 2, 4, 8, 12); the analysis dataset therefore comprised all randomized patients with complete data at every time point (*n* = 9 for cohort-level analyses; *n* = 3 per dose arm for arm-stratified analyses), and no imputation was required.

Prespecified analyses include the group-level comparisons of MADRS from baseline to week 4 (primary outcome) and of all continuous clinical or cognitive scores from baseline to each follow-up visit (secondary outcomes) using paired-samples *t*-tests. Post hoc and exploratory analyses include non-parametric sensitivity analyses of baseline-to-week-4 changes, linear mixed-effects models of temporal and illness-duration effects, between-arm effect-size estimation, and clinical-neuroimaging correlations.

Due to the small sample size (*n* = 9), the Wilcoxon signed-rank test was used as a non-parametric sensitivity analysis comparing the changes in the same six outcomes from baseline to week 4, with *P* values Benjamini-Hochberg FDR-corrected across the six scales. The parametric and non-parametric tests reached the same FDR-corrected significance conclusion (FDR *q* < 0.05) for five of six outcomes (MADRS, HAM-A, IDS-SR, DST and DSST). For PDQ-D, the paired-samples *t*-test reached significance (FDR *q* = 0.027) whereas the Wilcoxon signed-rank test did not (FDR *q* = 0.078), reflecting two patients (R001, R002) with score increases and one patient (R007) with an unchanged score at week 4; the within-group week 4 finding for PDQ-D should therefore be interpreted with caution (Extended Data Fig. 3f; Extended Data Table 3).

To examine change over the full 12-week follow-up and to identify prognostic covariates, linear mixed-effects models of the form score ∼ time + illness duration + (1 | subject) were fit separately for each of the six outcome measures (MADRS, HAM-A, IDS-SR, DST, DSST, PDQ-D), with time modeled as a categorical fixed effect (baseline, day 3, weeks 1, 2, 4, 8, 12), illness duration (years) included as a fixed-effect covariate without a time interaction, and a random intercept per patient. Models were estimated by restricted maximum likelihood (REML), with type III *F* tests for the time effect and Wald tests for the illness-duration coefficient; *P* values were corrected for multiple comparisons using the Benjamini-Hochberg FDR procedure, applied separately within each family of six scale-wise tests.

For all continuous outcomes, between-arm effect sizes are reported as mean differences with Welch 95% confidence intervals (CIs) and Hedges’ *g* with small-sample correction (Extended Data Table 1a). Binary MADRS response and remission rates at week 4 are reported per arm with Wilson 95% CIs, and between-arm effects on binary outcomes are reported as both absolute risk differences and relative risks with Wald 95% CIs (Extended Data Table 1b). Given the small per-arm sample size (*n* = 3), these between-arm estimates are reported for transparency of effect size and precision and are not intended to support formal between-arm hypothesis testing.

Neuroimaging data were analyzed in SPM using a full-factorial analysis of variance based on the general linear model. Associations between clinical outcomes and neuroimaging metrics were assessed primarily using Pearson correlation coefficients, with Spearman rank correlation coefficients computed as a non-parametric sensitivity analysis; since the sensitivity analysis did not alter the pattern of results, only the Pearson correlation coefficients are reported in the Results section.

### Diffusion MRI processing and tractography

Diffusion MRI data were acquired using a DTI diffusion scheme with 64 diffusion directions, with a resolution of 1.5 mm isotropic. Restricted diffusion was quantified using restricted diffusion imaging^41^. Diffusion data were reconstructed using generalized q-sampling imaging with a diffusion sampling length ratio of 1.25^42^. Deterministic fiber tractography was performed in DSI Studio using augmented tracking strategies to improve reproducibility^43,44^. All diffusion reconstruction and tractography analyses were performed using baseline diffusion MRI data. The anisotropy threshold was set to 0.05, the angular threshold to 70°, and the step size to the voxel spacing. A total of 5,000 seed points were placed. Topology-informed pruning (TIP) was applied for one iteration to remove false-positive fiber trajectories^45^. Analyses were conducted using DSI Studio^46^, Hou version (June 23, 2026).

For each patient, the individualized sgACC target was used as the seed region. All masks were transformed into the individual diffusion space using Advanced Normalization Tools (ANTs)^47^. The cingulum bundle (CB) was derived from the HCP-842 tractography atlas^48^. The mPFC mask comprised the bilateral A10m and A14m parcels from the Brainnetome Atlas^49^, in union with the mPFC regions showing significant functional-connectivity effects in this study. Separate sgACC-seeded tractography analyses were performed with streamlines required to traverse the cingulum bundle or terminate in the mPFC.

## Data availability

The raw imaging and clinical data are not publicly available due to patient privacy and institutional data-sharing policy but are available from the corresponding author on reasonable request.

## Code availability

Custom code used for statistical analyses is available at GitHub at https://github.com/fvz5058/trd-radiosurgery-sgacc. No other custom code or algorithms central to the conclusions of this paper were used; DPABI and DSI Studio are established, publicly available third-party software packages used as described in Methods.

## Acknowledgements

We thank the study participants and their families. We thank Y. Miao and J. Yang from Neuray Medical Technology Co. Ltd. for contributions to the initiation and funding of the study, and P. Nakrani from ZAP Surgical Systems, Inc. for manuscript consultation.

## Author contributions

A.Y., X.J., H.W., L.P., J.R.A. and G.W. conceived and designed the study. Y.Z., A.Y., X.J., Z.Z., X.Z., Z.Y., L.Z., H.W., L.P. and G.W. developed the study protocol and obtained institutional review board approval. Y.Z. recruited participants and obtained informed consent. Y.B., Z.Z. J.W., C.W. and L.P. performed the stereotactic radiosurgery target delineation and treatment planning. Y.B., J.W., C.W., X.L., B.Q., J.Z. and L.P. delivered the radiosurgical treatment. Y.Z acted as the investigator. Y.X., Q.Q., G.S. and Yuting W. acted as outcomes assessors and conducted clinical and cognitive assessments. A.Y. and H.H. acquired the neuroimaging data. A.Y., X.J., Z.Z., F.Z., B.W., Yun W. and H.W. performed neuroimaging preprocessing and analysis. F.Z., Yun W. and Q.M. performed the statistical analysis. Y.Z., F.Z., Yun W., Q.M. and H.W. prepared the data visualizations. Y.Z., X.J., F.Z., Yun W., Q.M., H.W. and J.R.A. wrote the first draft of the manuscript. All authors contributed to data interpretation and approved the final version for submission. G.W. is the guarantor of the study and had full access to all the data and takes responsibility for the integrity of the data and the accuracy of the data analysis.

## Extended Data

**Extended Data Fig. 1.**
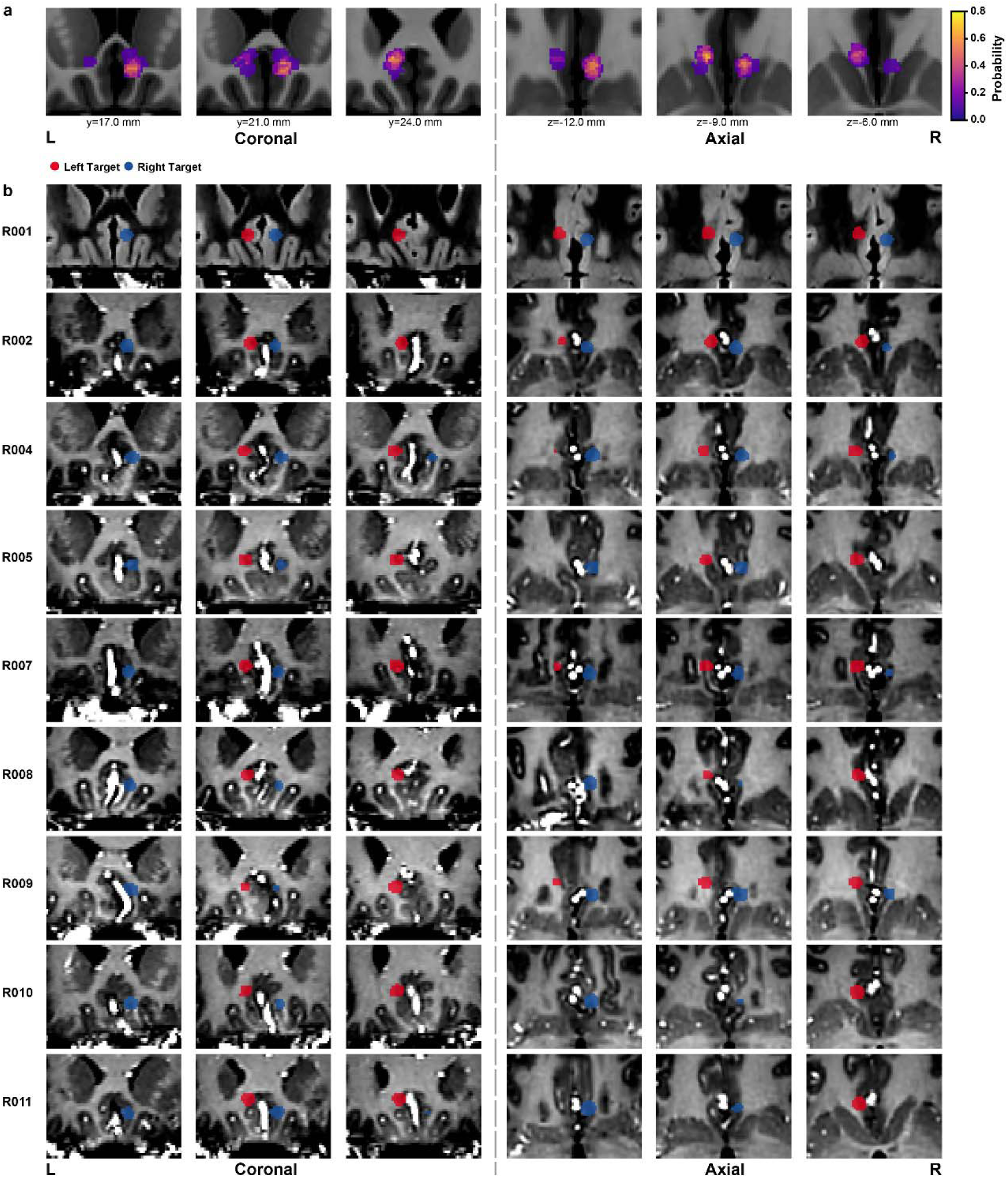
Bilateral sgACC target definition for stereotactic radiomodulation. Two-dimensional T1-weighted MR images showing the manually defined sgACC targets within the ZAP-X treatment planning system. **a**, Overall target distribution probability map in Montreal Neurological Institute (MNI) space, displayed in coronal and axial views with coordinates. **b**, Participant-specific sgACC targets in native space, displayed in coronal and axial views with coordinates; left- and right-hemisphere targets are shown in red and blue, respectively.

**Extended Data Fig. 2.**
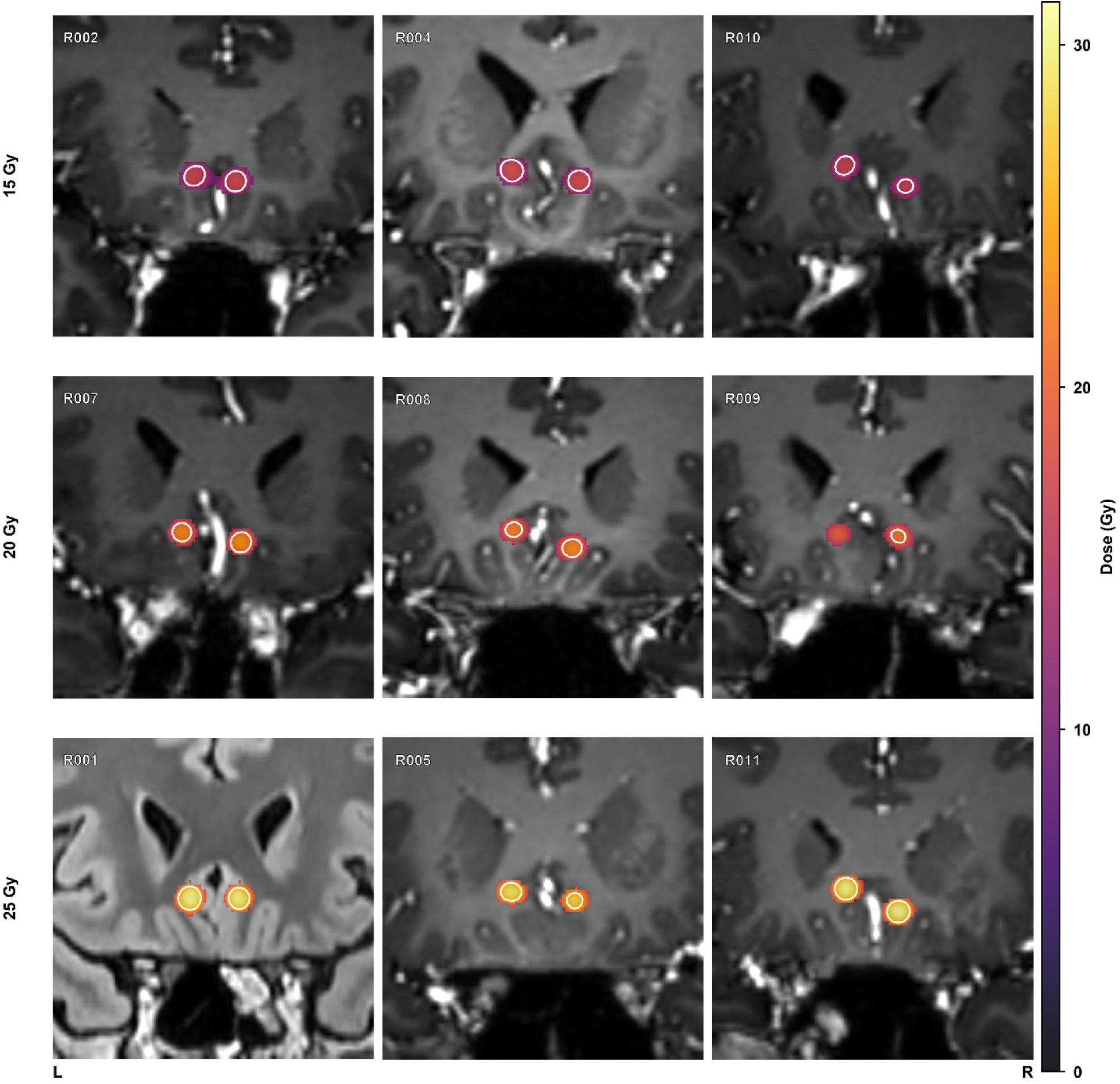
Dose distribution across the bilateral sgACC targets for all study participants. Coronal T1-weighted MR images depicting the delivered radiomodulation dose to the bilateral subgenual anterior cingulate cortex (sgACC) in nine participants, grouped by prescribed dose level (15 Gy, top row; 20 Gy, middle row; 25 Gy, bottom row). Each panel corresponds to an individual participant (identifier shown at upper left), with an enlarged inset providing a magnified view of the bilateral targets.

**Extended Data Table 1.**
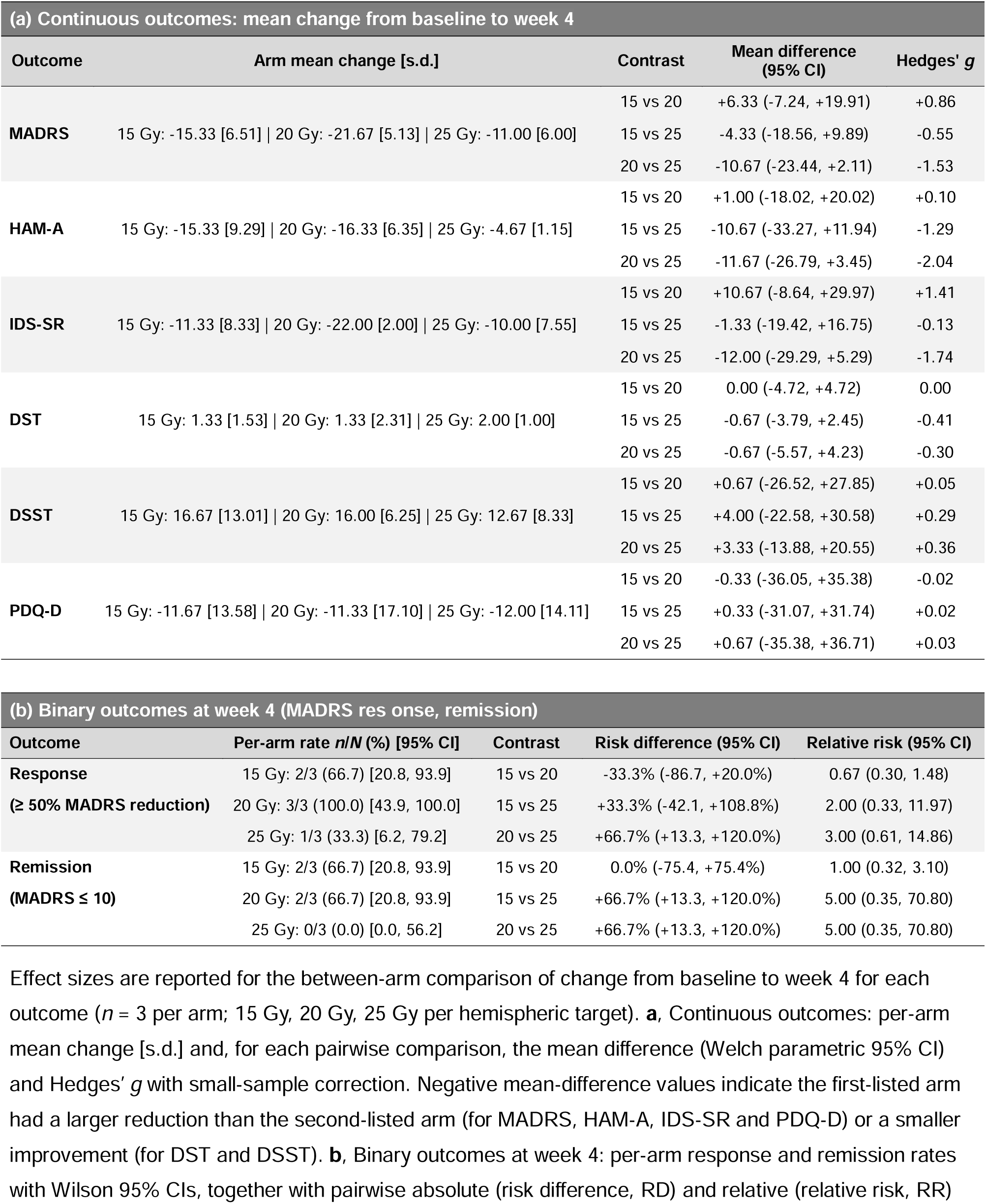
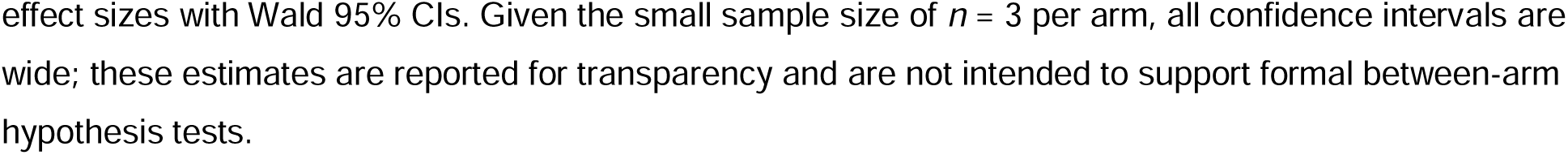
Between-arm effect sizes for the change from baseline to week 4 across primary and secondary outcomes.

**Extended Data Fig. 3.**
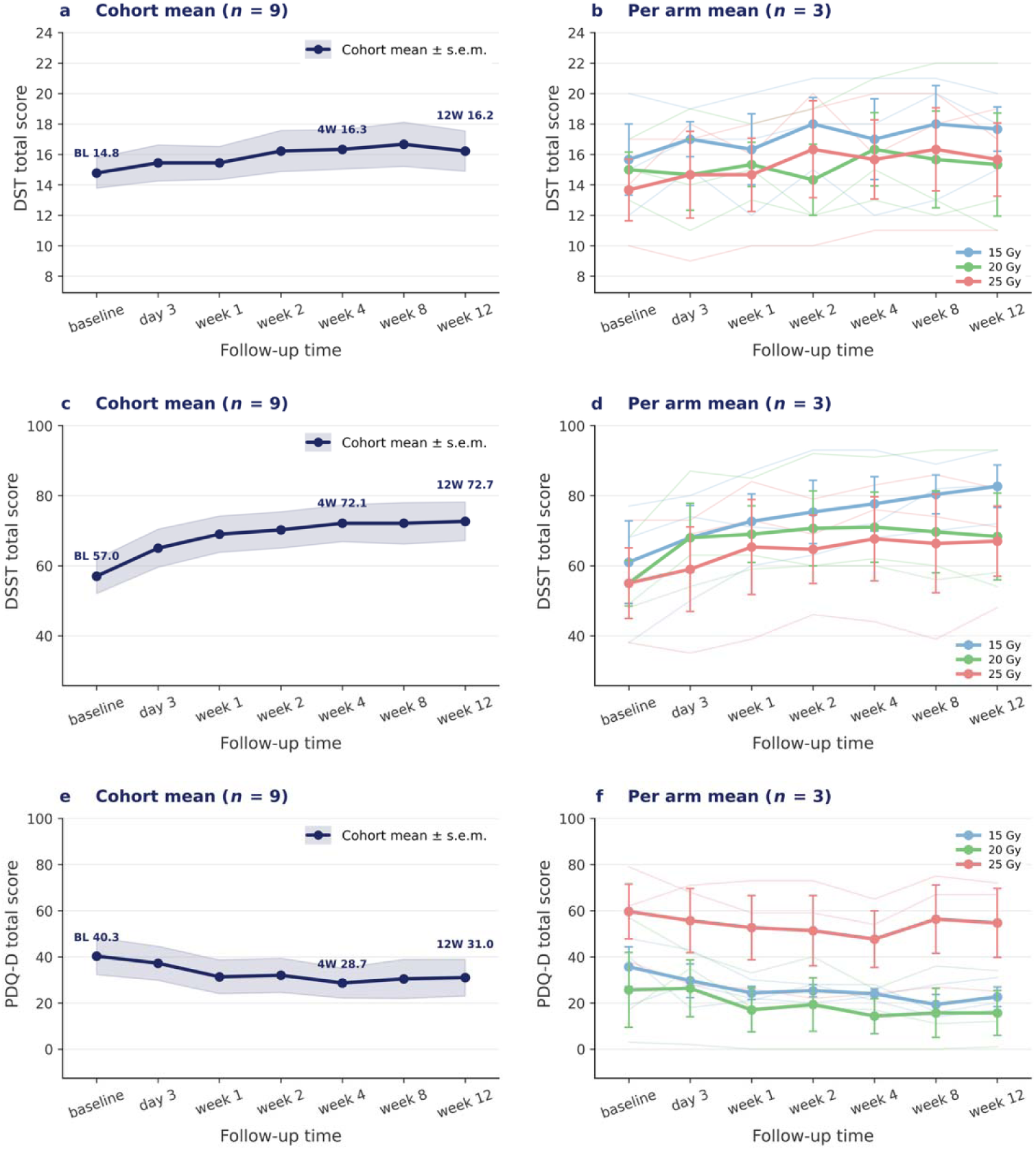
DST, DSST and PDQ-D total scores through the 12-week follow-up. **a**,**c**,**e**, Cohort mean (*n* = 9) ± s.e.m. at baseline, day 3 and weeks 1, 2, 4, 8 and 12; shading indicates the s.e.m. **b**,**d**,**f**, Per-arm means (*n* = 3 each) ± s.e.m. at 15 Gy (blue), 20 Gy (green) and 25 Gy (red) per hemispheric target; error bars indicate the s.e.m. and faint lines show individual participant trajectories. Clinician-administered cognitive performance improved on both measures: mean Digit Span Test score increased from 14.8 to 16.3 (group-mean improvement 10.5%; **a** and **b**), and mean Digit Symbol Substitution Test score increased from 57.0 to 72.1 (group-mean improvement 26.5%; **c** and **d**). Self-rated perceived cognitive deficits on the Perceived Deficits Questionnaire–Depression (PDQ-D) decreased from a baseline mean of 40.3 to 28.7 at week 4 (group-mean reduction 28.9%; **e**), with per-arm trajectories shown in **f**.

**Extended Data Table 2.**
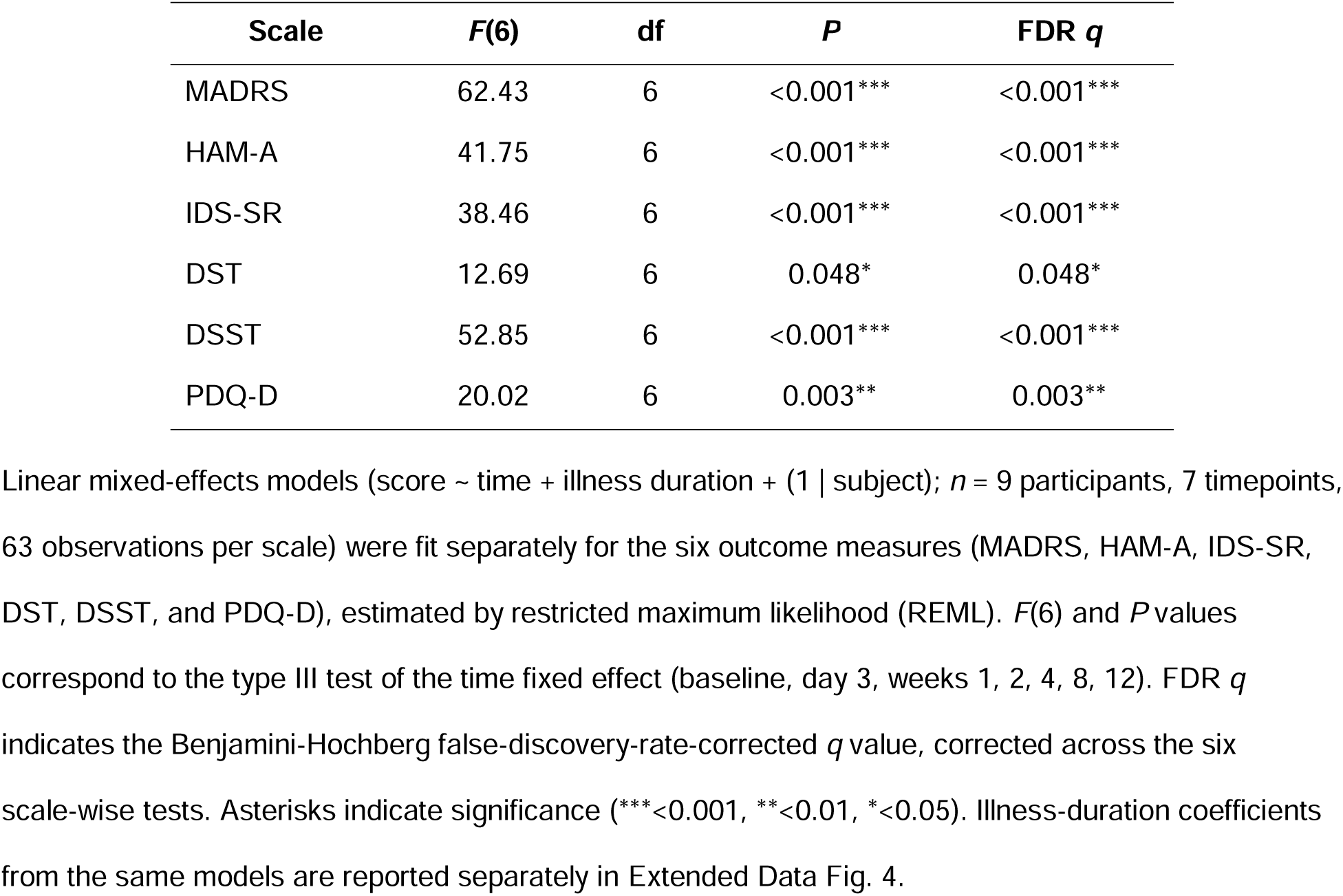
Time effects from linear mixed-effects models predicting outcome scores over 12 weeks.

| Scale | *F*(6) | df | *P* | FDR *q* |
| --- | --- | --- | --- | --- |
| MADRS | 62.43 | 6 | <0.001*** | <0.001*** |
| HAM-A | 41.75 | 6 | <0.001*** | <0.001*** |
| IDS-SR | 38.46 | 6 | <0.001*** | <0.001*** |
| DST | 12.69 | 6 | 0.048* | 0.048* |
| DSST | 52.85 | 6 | <0.001*** | <0.001*** |
| PDQ-D | 20.02 | 6 | 0.003** | 0.003** |

**Extended Data Table 3.**
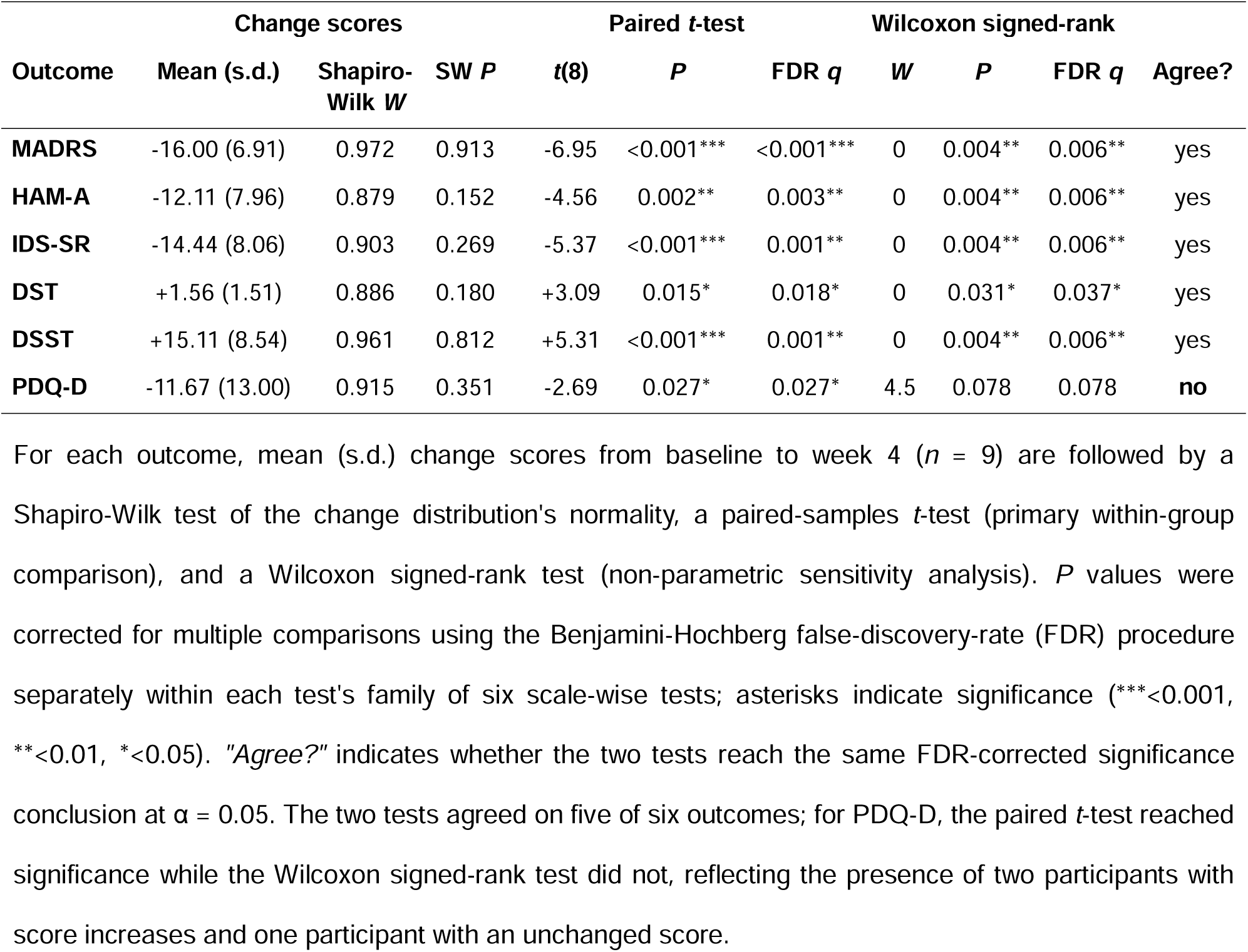
Sensitivity analysis: paired-samples *t*-test versus Wilcoxon signed-rank test for the change in clinical outcomes from baseline to week 4.

**Extended Data Fig. 4.**
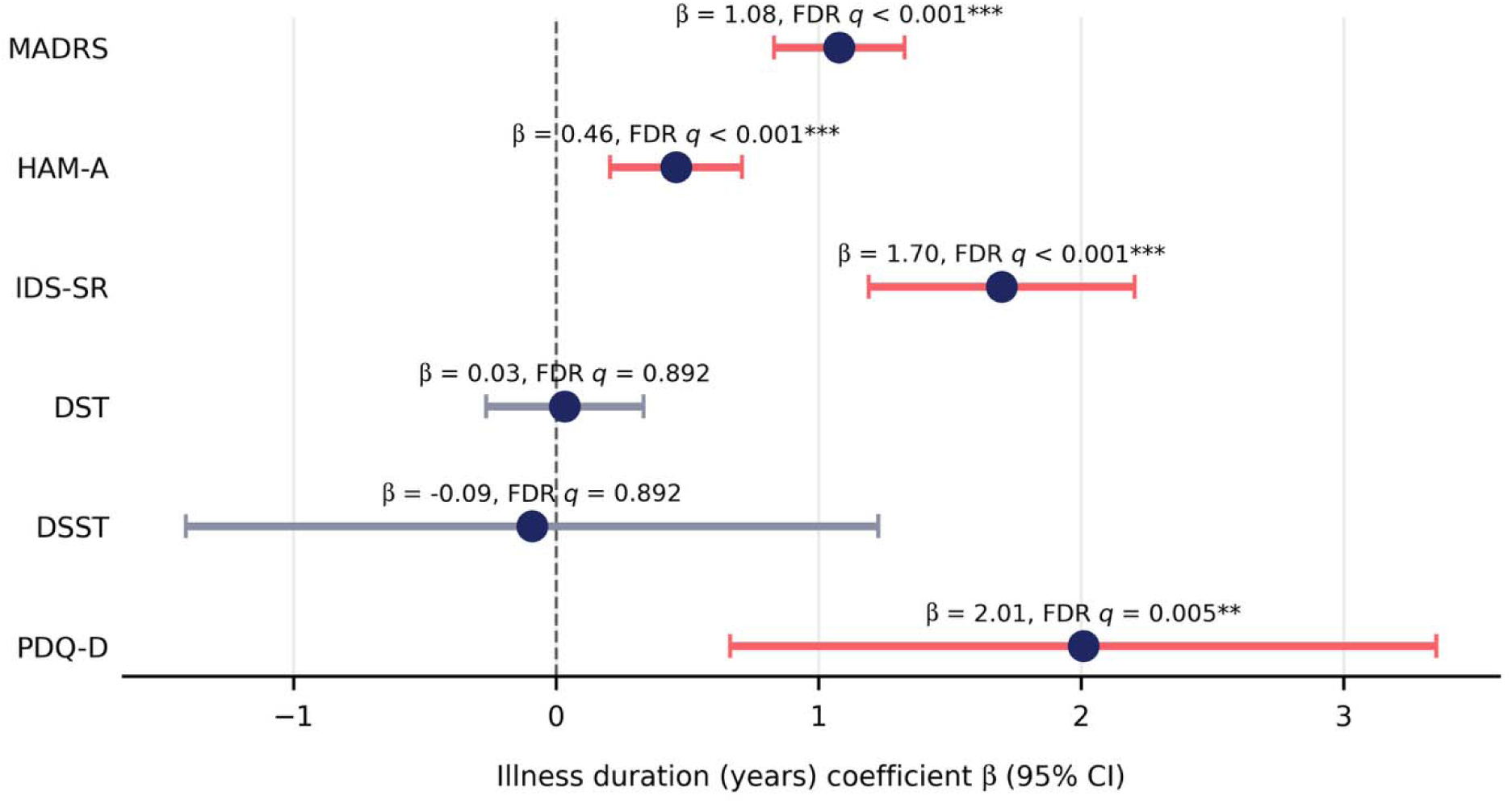
Fixed-effect estimates of illness duration on symptom and cognitive outcomes. Forest plot of illness-duration coefficients (β, points) and 95% confidence intervals (error bars) from linear mixed-effects models (score ∼ time + illness duration + (1 | subject); *n* = 9) fit separately for each outcome measure. Positive β indicates that longer illness duration (years) is associated with higher scores on that measure. Coefficients for MADRS, HAM-A, IDS-SR and PDQ-D are shown in red (FDR *q* < 0.05); coefficients for DST and DSST are shown in grey (not significant after FDR correction). FDR *q* values above each estimate show the Benjamini-Hochberg FDR-corrected *q* value for that scale’s illness-duration coefficient, corrected across the six scales. Asterisks indicate significance (***<0.001, **<0.01, *<0.05). The dashed vertical line indicates no effect (β = 0).

**Extended Data Fig. 5.**
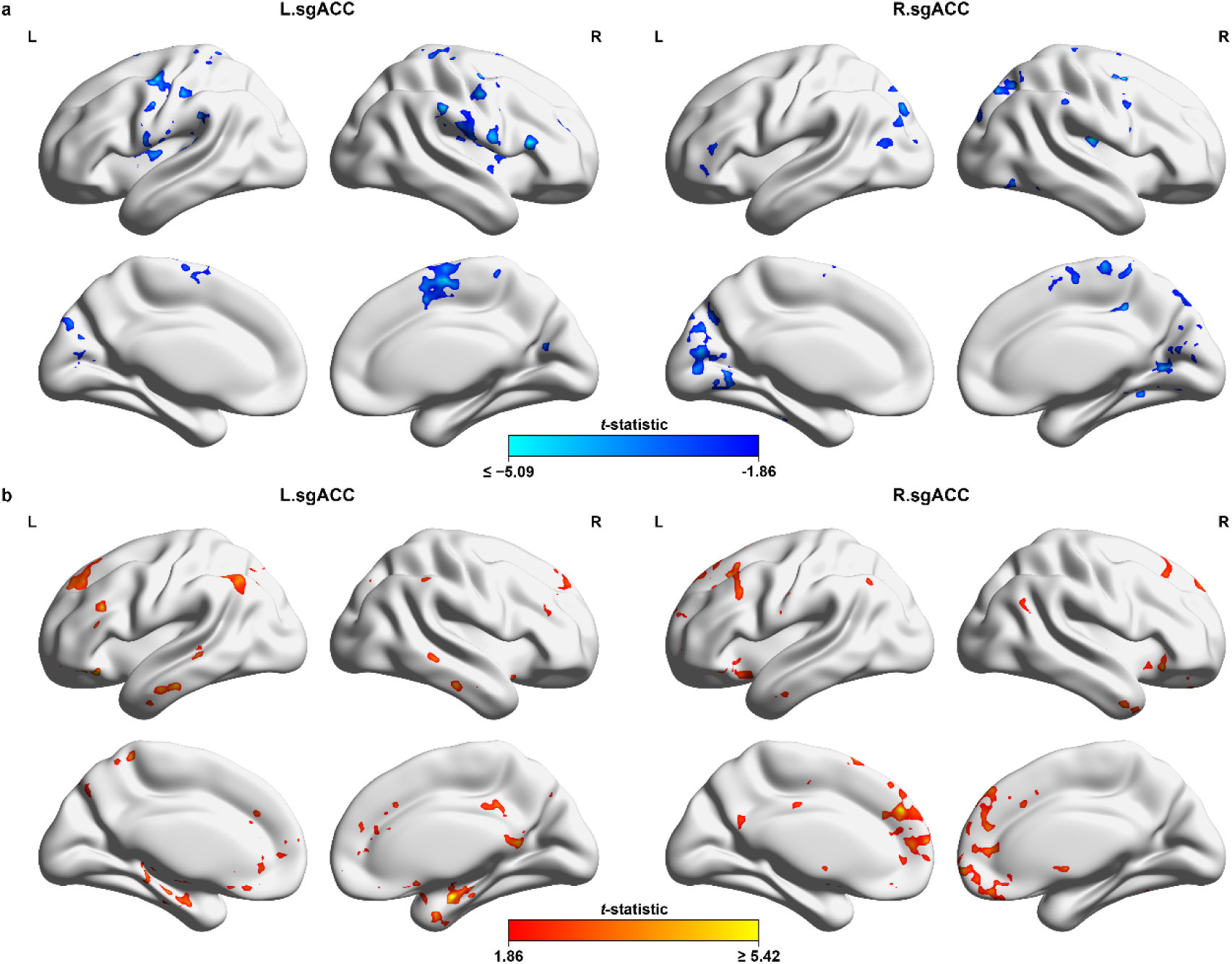
Functional connectivity changes after sgACC radiomodulation. **a**, Using a liberal statistical threshold (voxel-level *P* < 0.05, cluster extent > 100 voxels, uncorrected), target-based functional connectivity analyses revealed stimulation-related decreases in connectivity across several brain regions. Relative to the left sgACC (L.sgACC), the right sgACC (R.sgACC) also exhibited a trend toward reduced connectivity with the precentral gyrus (PreCG), although this effect was less pronounced. **b**, Using a liberal statistical threshold (voxel-level *P* < 0.05, cluster extent > 100 voxels, uncorrected), target-based functional connectivity analyses revealed stimulation-related increases in connectivity across several brain regions. Relative to the right sgACC (R.sgACC), the left sgACC (L.sgACC) also exhibited a trend toward increased connectivity with the medial prefrontal cortex (mPFC), although this effect was less pronounced. Color scales were truncated at the 99^th^ percentile; untruncated peak *t*-values for the left/right sgACC maps were -11.19/-10.19 in **a** and 15.08/12.69 in **b**.

